# The influence of different exercise types on elderly depressed patients: protocol for an umbrella review

**DOI:** 10.1101/2022.06.26.22276887

**Authors:** Jie Liu, Shu-qing Gao, Lian-cheng Zhang

## Abstract

**Introduction:** At present, there are a large number of studies confirming that exercise can improve the depression of the elderly, but there is no research to compare the effects of different exercise programs. Therefore, the purpose of this article is to compare the effects of different exercise programs on the mood improvement of elderly people with depression, and select suitable exercise programs., In order to provide guidance and suggestions for clinical practice.

**Methods and analysis:** Document retrieval was carried out in 7 databases including CNKI, ebsco, PsycARTICLES, PsycINFO, PubMed, Cochrane Library and Google Scholar. The retrieval time was from January 1, 2000 to May 1, 2022.

**Results:** Obtain the exact amount of effect of different exercise programs on the mood improvement of elderly depression patients. This study will provide more convincing evidence for the effect of physical exercise on improving the mood of the elderly with depression. In addition, by comparing the effects of different exercise programs, the best program for the depression of elderly people with depression can be obtained, and some specific exercise suggestions can be provided for clinical practice, so as to promote the successful aging of the elderly.

**Ethics and dissemination:** As this research is a systematic review of published literature, ethical approval is not required. The results will be reported according to the latest guidelines for Preferred Reporting Items for Systematic Reviews and Meta-Analyses 2020 statement, and will be submitted to a peer-reviewed journal.

**PROSPERO registration number:** CRD42021250602.

**Strengths and limitations of this study:**

- The planned systematic review will systematically report on the improvement of different types of exercise in elderly patients with depression.
- This protocol describes a comprehensive search strategy and eligibility criteria, which have no geographical restriction.
- The systematic review might be limited by the presence of selection and/or attrition bias inherent in some of the selected studies.
- Find the best exercise prescription for older adults with depression
- As with many reviews, the results may yield significant heterogeneity among the included studies.

## 1 Introduction

Depression is one of the most common mental health diseases in the world today. The World Health Organization survey found that there are approximately 322 million cases of depression in the world, accounting for 4.4% of the world’s population. Many of them are among the elderly. Depression has become a major disabling disease in the world. Rise to the world’s first burden of disease (Friedrich, 2017). Although the treatment of depression is diverse, most of them are still limited to medication and psychotherapy (Lopresti, 2019). The study found that although more than 30% of elderly depression patients have undergone multiple treatment trials, they have not yet been relieved (Friedrich, 2017). Therefore, it is urgent to develop new treatment methods to improve or treat depression. A study by Lopresti (2019) found that the elderly who regularly participate in exercise are often at relatively low risk of depression; in addition, the mental health benefits of exercise are more obvious in people with anxiety and depression. A Mate analysis of 42,264 people found that exercise can significantly improve anxiety (Wegner et al., 2014). In fact, exercise has been specifically used as a treatment for depression (Toups et al., 2017). Studies have shown that compared with people who engage in less or no exercise, people who exercise 2 to 3 times a week have a lower risk of depression, anger, and depression (Kruk et al., 2019). Therefore, in view of the psychological and physical health benefits of physical exercise, more and more researchers pay attention to physical exercise as an alternative or auxiliary treatment for improving depression. This shows that regular exercise is essential for elderly depressed patients. In addition, physical exercise, an alternative medication intervention, is often used to cope with widespread depression and has positive results (Bertisch, 2009; Cramer, 2013; Sharma & Haider, 2013; Tsang, 2008). So is the positive effect of physical exercise on the depressive symptoms of the elderly equally significant? Are the effects of different types of physical exercises different?

At present, physical exercise is widely used as exercise therapy for elderly depression patients, and there are many related systematic reviews. However, so far, the overall quality of research on the positive effects of different exercise programs on elderly depression patients is uneven and the conclusions are not consistent. For example, some studies have shown that Laughter Yoga significantly reduces elderly depression compared with non-exercise control groups. The depression scores of women with severe symptoms, but no difference was found between the yoga group and the aerobic exercise group (Shahidi et al., 2011). Two other studies have found that aerobic exercise is as effective as sertraline (a commonly used antidepressant) in the treatment of depression in the elderly (Brenes et al., 2007), while Tai Chi is more effective in treating depression in the elderly than alone The use of antidepressants is more effective (Lavretsky et al., 2011), which leads to the conclusion that Tai Chi therapy is more effective than aerobic therapy. Some scholars believe that the elderly have good physical responses to resistance exercises, and that psychological improvements may occur at the same time as physical improvements. The effects of resistance exercises are significantly better than all other types of activities. (Arent, 2000). Conflicting results may interfere with the formulation of exercise prescriptions for elderly depression patients, and may also cause confusion for patients.

Literature review review (review of reviews) is a special research paradigm in evidence-based medicine research. It usually appears when the research on related topics is relatively mature and it is necessary to conduct a higher-level summary of the current research (Lou Hu, 2018). At present, the relationship between physical exercise and the elderly depressed population has not yet appeared to be a higher-level summary of existing reviews. Therefore, this article will conduct an umbrella review method based on the meta-analysis of the RCT experiment with elderly depression patients as the research object, which is to collect high-quality evidence from the relevant meta-analysis or review literature., The effect of physical exercise on improving the mood of elderly depression is systematically evaluated again, and higher-level high-quality conclusions are obtained. This conclusion can provide references for theoretical research, and more importantly, it can provide clearer information for clinical practitioners. Views, in order to formulate detailed exercise prescriptions for improving the mood of the elderly depressed people, and contribute to the promotion of successful aging in our country.

## 2 Method

The network meta-analysis extension for the Preferred Reporting Items for Systematic Reviews and Meta-Analyses (PRISMA-NMA) guideline (Hutton,2015) and the Cochrane Intervention Review that Compares Multiple Interventions (Cochrane,2014) each provided further support in guiding this review. Guidelines specific for geriatric meta-analyses (Shenkin,2017) were consulted to identify baseline characteristics, equity considerations, inclusion/exclusion criteria, known confounders, and potentially important effect modifiers.

### 2.1 Search strategy

First of all, we will start searching in 7 databases of CNKI, ebsco, PsycARTICLES, PsycINFO, PubMed, Cochrane Library and Google Scholar from January 1, 2000. The search language is English or Chinese, and the search strategy will be based on subject terms. And free words. PubMed’s search strategy is shown in Table 1.

**Table 1.** PubMed search strategy

| serial number | search term |
| --- | --- |
| #1 | exercise [Title/Abstract] |
| #2 | Physical activity [Title/Abstract] |
| #3 | aerobic [Title/Abstract] |
| #4 | resistance [Title/Abstract] |
| #5 | Mind-body [Title/Abstract] |
| #6 | #1 OR #2 OR #3 OR #4 OR #5 |
| #7 | systematic review [Publication Type] |
| #8 | overview [Publication Type] |
| #9 | meta-analysis [Publication Type] |
| #10 | pooling analysis [Publication Type] |
| #11 | #7 OR #8 OR #9 OR #10 |
| #12 | Elder |
| #13 | old |
| #14 | #12 OR #13 |
| #15 | depression |
| #16 | #6 AND #11 AND #14 AND #15 |

### 2.2 Inclusion criteria

The criteria for inclusion in the literature are mainly judged in four aspects: participants, intervention methods, outcome indicators and research design types. Participants are required to be elderly people over 55 years of age suffering from depression. The intervention method is to use physical exercise to intervene. The outcome indicators require clear data on the participants’ depression scale before and after the intervention, and the type of study is required. It is a review study. It will be excluded when the intervention condition uses a multi-component treatment, including non-exercise components related to the exercise condition. There are no restrictions on gender, age, race, nationality, occupation, marital status, economic status, education level, etc.

### 2.3 Intervention and comparison

We will take physical exercise as an intervention measure, and compare it with conventional treatment, conventional nursing, and health education. There are no restrictions on the frequency, intensity, or duration of physical exercise.

### 2.4 Results measurement

We will include all systematic reviews and meta-analyses based on randomized controlled trials. The main outcome indicator is the score on the depression scale. Secondary outcomes will be positive emotions, anxiety, self-efficacy, etc. There are no restrictions on secondary results. If other results are reported in related studies, they will also be considered for inclusion in the outcome measurement.

## 3 Data collection and evaluation

### 3.1 Research options

The researcher imported all the documents retrieved to meet the standards into the document management tool note express to delete duplicate documents. Then, the imported documents are manually screened. First, two reviewers are responsible for half of the documents, and the titles and abstracts are independently screened according to the inclusion criteria, and the documents that meet this topic are selected. Then, the two exchanged the content of the screening, cross-checked, and the third reviewer resolved their differences. Second, the same two reviewers will independently review the full text articles left after the first screening. The results were cross-checked again, and the differences and the same third-party reviewer resolved differences. The details of the selection process are given in the systematic review and meta-analysis (PRISMA) flowchart (Figure 1).

**Figure 1.**
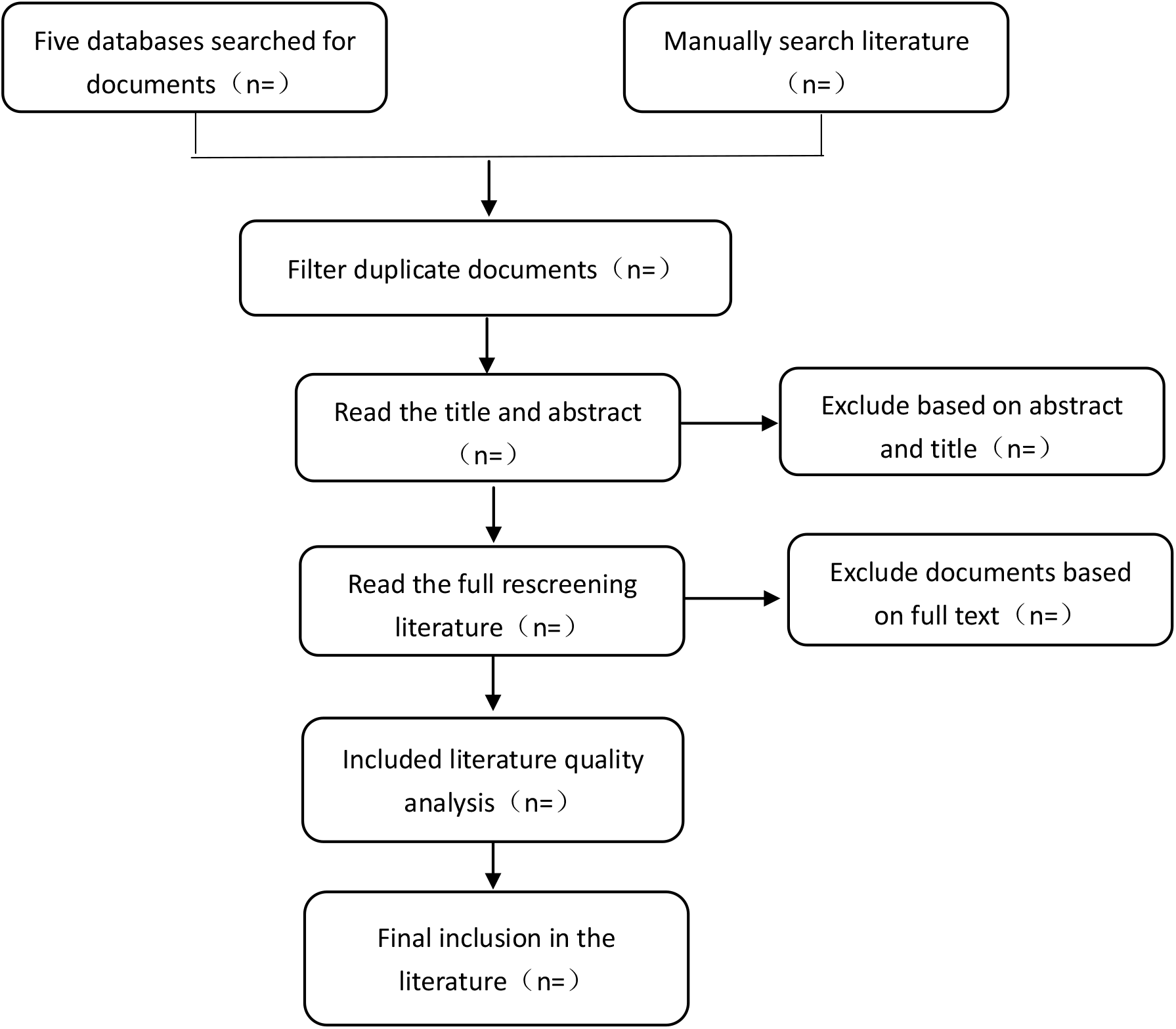
Flow chart of the screening process

### 3.2 Data extraction and management

Two reviewers will independently extract data according to the preset data extraction table, and cross-check the extracted data to reach a consensus. The data extraction table mainly includes the author’s name, publication year, country, sample size, disease category, participants, intervention, comparison, results, etc. If the required data are insufficient or missing, the author of the relevant systematic review will be contacted.

### 3.3 Quality assessment

The same two reviewers will independently assess the quality of the included reviews included in the measurement tool AMSTAR-2 declared by PRISMA (the preferred reporting project for systematic reviews and meta-analysis), and the same third reviewer will discrepancies will resolve it.

### 3.4 Report quality assessment

The “PRISMA Statement” will be used to assess the quality of the included literature. The “PRISMA Statement” is a 27-item checklist, which is mainly aimed at the systematic evaluation of randomized controlled trials. The scoring principles are as follows: each item is scored 1 (reported and sufficient), 0.5 (reported and insufficient) or 0 (unreported). The total score ranges from 0 to 27 points. A score of less than 15 indicates a serious reported defect, a score of 15 to 21 indicates a partial report of a defect, and a score of 22 to 27 indicates a relatively complete report.

## 4 Data analysis

A narrative synthesis of the system review will be conducted. The summary of the research characteristics, report and method quality assessment and the main conclusions will be given in detailed tables. Where appropriate and feasible, quantitative analysis will be conducted on the basis of available data.

## 5 Discussion

A large number of systematic reviews have shown that physical exercise can effectively improve the health of elderly patients with depression. However, there are still great uncertainties in the quality of these pieces of evidence, which hinders the clinical application of these findings. Because of it is necessary to summarize and evaluate the existing evidence. This one is the first umbrella evaluation to evaluate the impact of physical exercise on senile depression. We hope that when using physical and mental exercise as exercise therapy in elderly depression patients, it can provide clinicians and patients with more convincing evidence. However, this umbrella evaluation has some limitations. Because the frequency, intensity, and duration of exercise differ between studies, there is a significant clinical heterogeneity, which may make it difficult to quantitatively analyze the effectiveness of physical exercise.

## Data Availability

All data produced in the present study are available upon reasonable request to the authors. All data produced in the present work are contained in the manuscript

## Contributors

Gao and Zhang designed the study. Liu registered the protocol with the PROSPERO database. Gao and Liu drafted the manuscript, which was critically revised by Zhang. All authors agreed on the final manuscript for submission.

## Patient and public involvement

Patients and/or the public were not involved in the design, or conduct, or reporting, or dissemination plans of this research.

Patient consent for publication Not applicable.

